# A study of pelvic organs’ sizes and findings in transvaginal sonography

**DOI:** 10.1101/2025.01.11.25320401

**Authors:** Farzana Rahman Adrita, Azmal Kabir Sarker, Sanjida Akter, Sabuz Paul

## Abstract

**Purpose:** This retrospective cross-sectional study evaluated the correlation among pelvic organs’ size, correlation of sizes with age, the change of uterine and cervical sizes in presence and absence of pathologies, and the pattern of ovarian volume asymmetry in the absence of pathology.

**Patients and methods:** Measurement of pelvic organs from all consecutive premenopausal, non-pregnant patients who underwent TVS for gynecologic workup were included in the analysis. The correlation of the sizes, and variation of sizes among a finding-based category was checked.

**Results:** Data from 43 women with mean (±SD) age of 33.4±8.7 years were included. Among the finding-based categories, bulky uteri in comparison to normal uteri were more likely to harbor myoma or adenomyosis (Risk ratio 2.07, p < 0.01). Positive correlations were found between, antero-posterior (AP) diameter of uterus and age (R 0.4, p < 0.01), AP diameter of uterus and its length (R 0.7, p < 0.01), AP diameter of uterus and cervix (R 0.44, p < 0.01), length of uterus and that of cervix (R 0.37, p < 0.05), AP diameter of cervix and it length (R 0.47, p < 0.01) and pair of otherwise healthy ovaries (R 0.53, p < 0.01). This dataset suggests that the presence of pathologies may not always be associated with uterine enlargement (p > 0.05), but the presence of a pathology is associated with enlargement of uterine cervical AP diameter (p < 0.01) as well as its length (p < 0.05). Among the healthy pairs of ovaries, the right one was larger than the left with a significant difference between the mean volumes (p < 0.05), in 70.4% pairs.

**Conclusions:** Overall, the findings from this small series are expected to add factual evidence to our existing concepts and help to shape some future research questions.

## INTRODUCTION

Uterine and ovarian pathologies are common conditions that affect women of reproductive age, often leading to clinical symptoms such as abnormal bleeding, pelvic pain, and infertility (1). Uterine myomas and adenomyosis are among the most frequently diagnosed structural abnormalities and major contributors for subfertility (2). With the age standardized incidence rate of myoma, Bangladesh belongs to the tiar of countries with the second highest global estimated annual percentage increase (3). Although the incidence of adenomyosis in the population is unknown, estimated prevalence of adenomyosis is up to 62% from the hysterectomy specimens (4) and up to 21% from diagnostic ultrasound scans (5). Additionally, the ovarian volume has implications in the management of adolescent abnormalities, polycystic ovarian syndrome, assessment of ovarian reserve, prediction of response to superovulation, and ovarian cancer. (6).

Transvaginal sonography (TVS) with measurement of uterus, cervix and ovaries is a major clinical tool for evaluation of pelvic organs for the management of gynecological and obstetric conditions. In the absence of a lesion, the size of uterus and cervix as well as the volumes of ovaries are reportedly associated with age, being at pre or perimenopausal state, anthropometric parameters, body composition parameters, fertility, parity, circulatory hormone levels, and use of hormonal contraceptive (7, 8).

This retrospective cross-sectional study was conducted to evaluate the correlation among pelvic organs’ size, correlation of sizes with age, the change of uterine and cervical sizes in presence and absence of pathologies, and the pattern of ovarian volume asymmetry in the absence of pathology.

## PATIENTS AND METHODS

Measurement of pelvic organs from all consecutive premenopausal, non-pregnant patients were included in the analysis. The parameters included in the analyses were length and fundal antero-posterior (AP) diameter of uterus, length and AP diameter of the cervix and volume of each ovary. Correlations of the parameters from uterus, cervix and were checked against the patients’ age. Then correlation was checked within the various parameters of uterus, cervix and ovaries, as well as between otherwise healthy pairs of ovaries. The significant results are shown. Furthermore, the findings from pelvic organs were categorized (according to box 1) to observe the change of uterine and cervical size among those categories, and to observe the variation of sizes between otherwise healthy pair of ovaries.

All statistical tests were done on R. Correlation plots were generated using the “ggplot2” and the box-violin plots were generated using the “ggstatsplot” functions. The waterfall plot was generated using Microsoft Excel.

### Box 1

Categories of findings from uterus, cervix and ovaries

Box 1: Categories of findings from uterus, cervix and ovaries
| Findings from uterus | Findings from cervix | Findings from ovary |
| --- | --- | --- |
| 1.Normal in size and appearance | 1. Normal in size and appearance | 1. Contain developing follicles |
| 2.Normal in size with other pathology | 2. Broad cervix without visible lesion | 2. Dominant follicles |
| 3.Bulky uterus with or without other pathology | 3. Any size with nabothian cyst or myoma etc | 3. Scanty to no follicles |
|  |  | 4. Polycystic appearance |
|  |  | 5. Mass, multilocular cyst or chocolate cyst |

## RESULTS

Data from 43 women with mean (±SD) age of 33.4±8.7 years were available for the analyses. The summary of findings among these women and are given in table 1. Apart from the 13 morphologically normal uterus, uterine myomas were found in four patients with normal sized uterus and non-homogeneous myometrium, the surrogate for adenomyosis, was found in seven patients with normal sized uterus. Among the patients who had bulky uterus, six had myomas, 12 had non-homogeneous myometrium and one had no visible structural pathology. Thus, bulky uteri in comparison to normal uteri were more likely to be associated with myoma or adenomyosis (Risk ratio 2.07, Fisher exact p = 0.0008 and Yates Chi-square 9.43, p = 0.002).

**Table 1.**
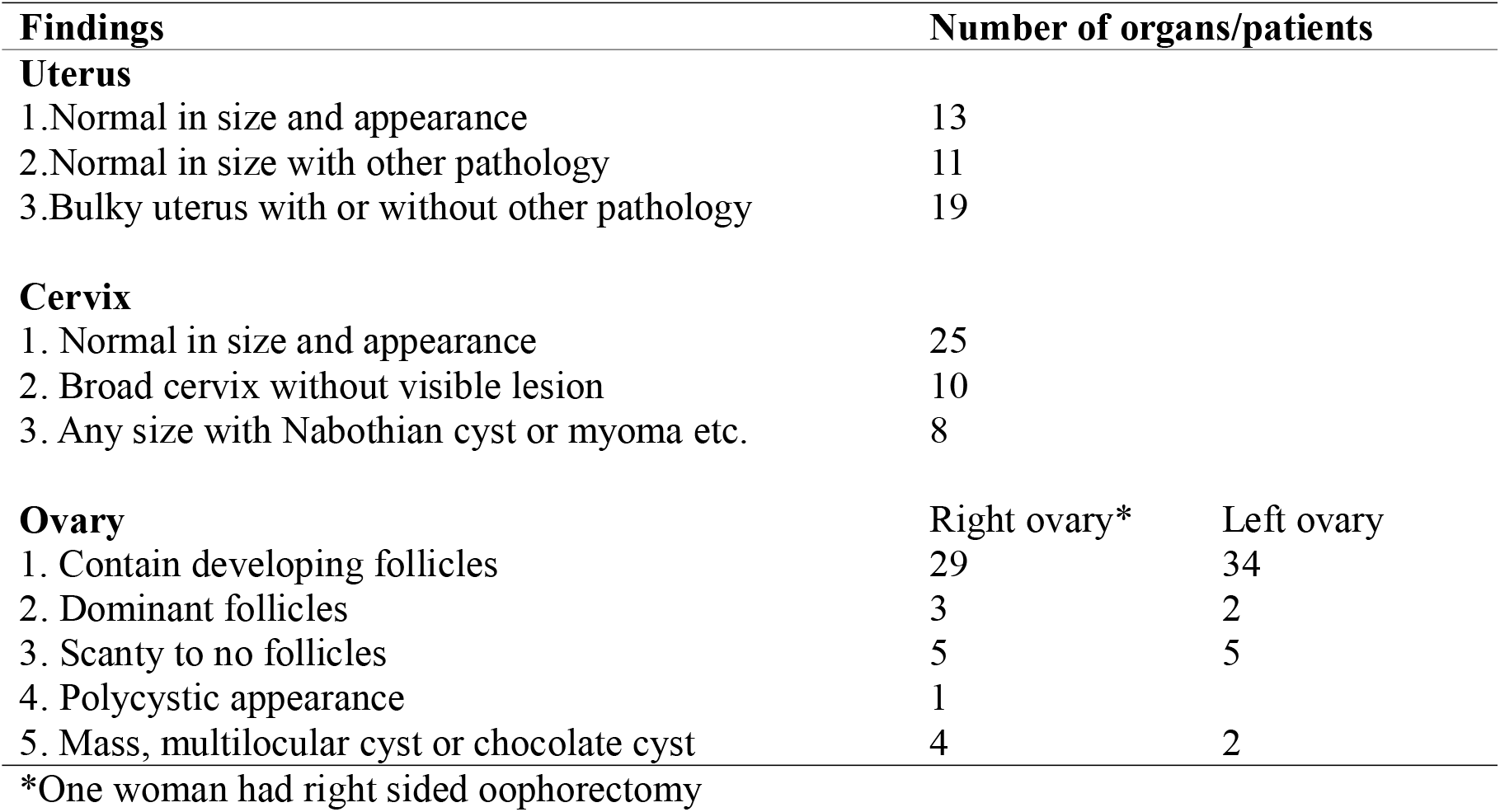
Summary of findings in the study subjects (n=43)

Moderately positive (Pearson’s R = 0.4) but significant (p = 0.07) correlation was found between AP diameter of uterus and patients’ age (Figure 1a). Among the intra-parameters’ correlation (figure 1b, 1c, 1d and 1e), significant correlation was found between uterine AP diameter and uterine length (R = 0.73, p < 0.001), between AP diameters of uterus and cervix (R = 0.44, p = 0.003), between length of uterus and cervix (R = 0.37, p = 0.014) and between length and AP diameter of cervix (R = 0.47, p = 0.001).

**Figure 1.**
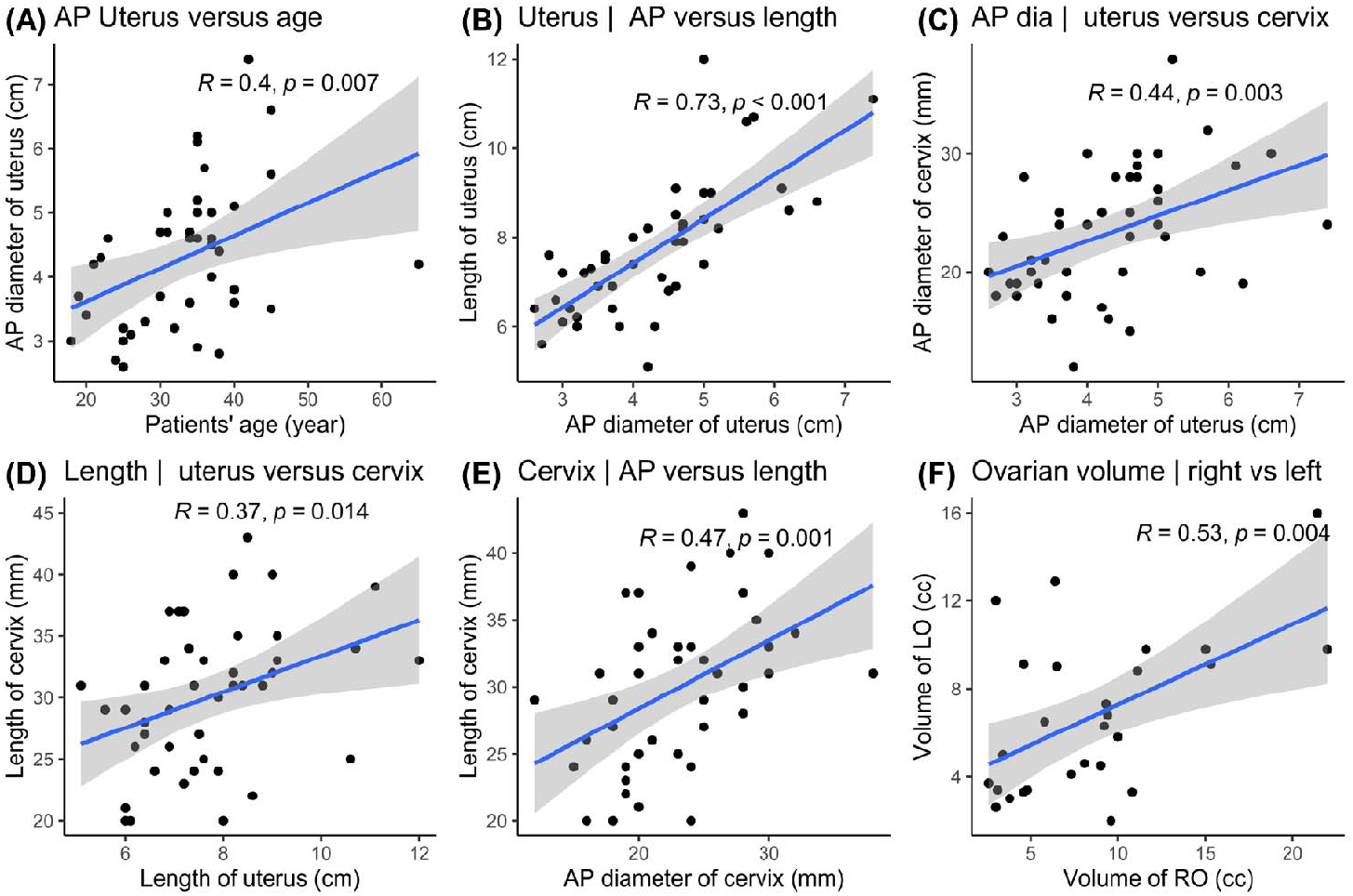
Scatterplots showing correlations between patients’ age and AP diameter of uterus (A), AP diameter and length of uterus (B), AP diameter uterus and AP diameter of cervix (C), Length of uterus and length of cervix (D), AP diameter of cervix and length of uterus (E), volume of ovaries (n = 27) containing developing follicles without any other feature (F).

The distribution of uterine AP diameters and uterine lengths across the three categories of uterine findings (figure 2a and 2b) shows, the mean of categories 1 and 2 weren’t significantly different, meaning the uterine pathology may not always be associated with uterine enlargement. The distribution of uterine cervical AP diameters and lengths across the three categories of cervical findings (figure 3a and 3b) shows, the mean of categories 1 and 2 were significantly different, meaning the cervical pathology may always be associated with an enlargement or broadening.

**Figure 2.**
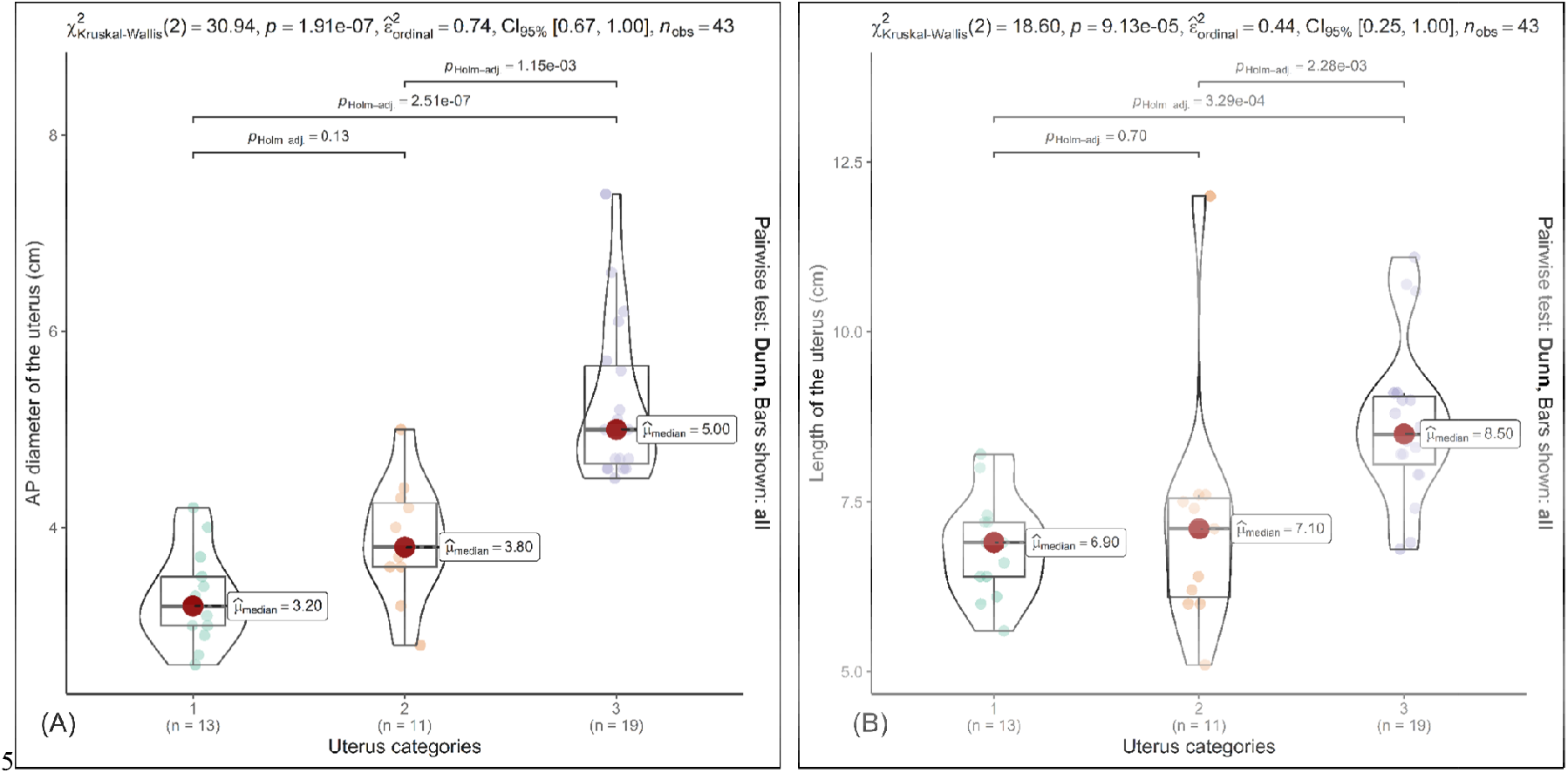
Box-violin plots show distribution of AP diameter of the uterus across the categories (a), and length of the uterus across the categories (b). The mean AP diameter and mean length of bulky uteri (category 3) was significantly different from with those with normal size with or without any associated pathology (p < 0.01) whereas the mean diameter and length of the categories 1 and 2 weren’t significantly different. This means that the presence of pathologies may not always be associated with uterine enlargement

**Figure 3.**
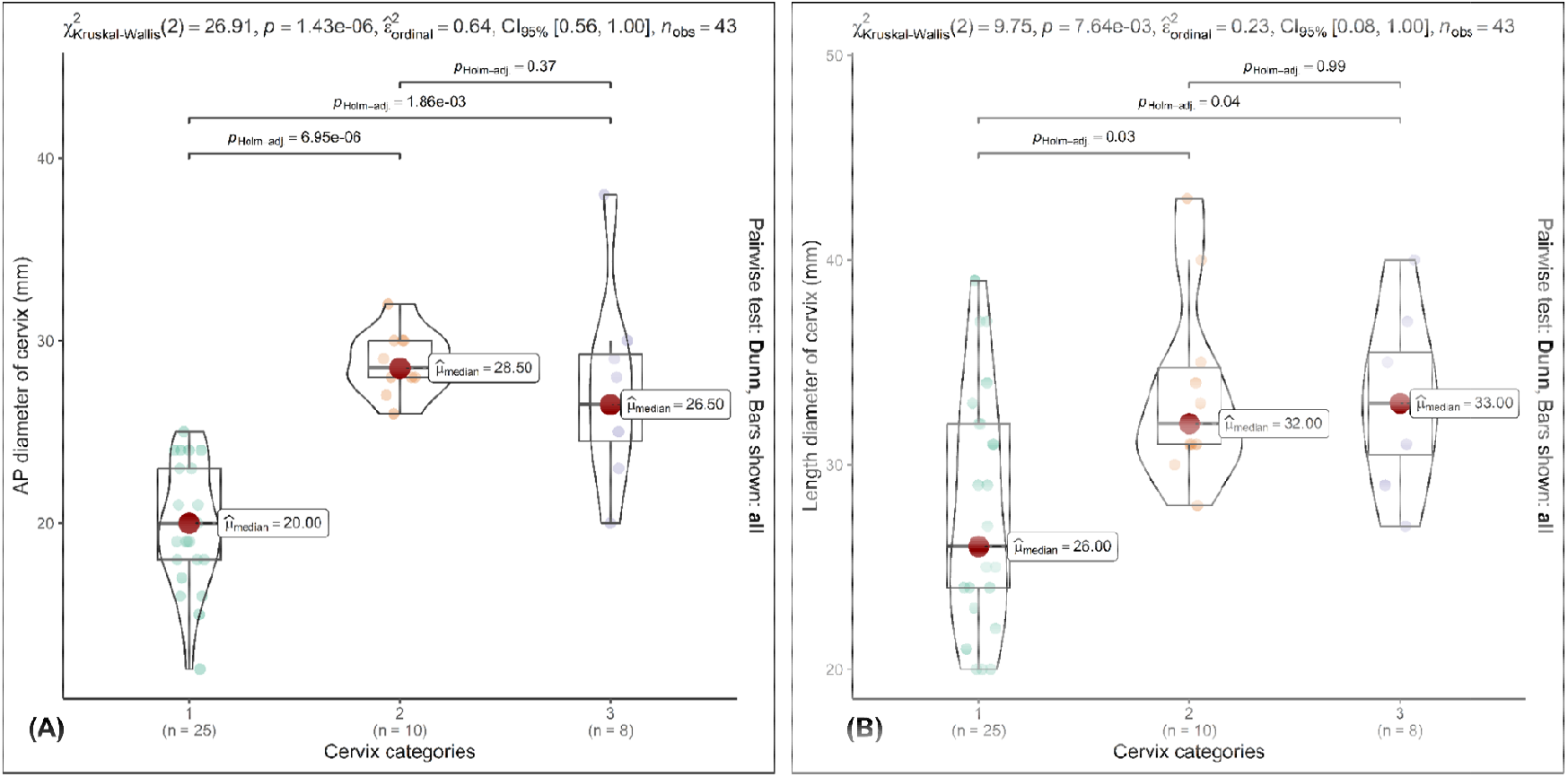
Box-violin plots show distribution of AP diameter of the uterine cervices across the categories (a), and length of the uterine cervices across the categories (b). The mean AP diameter and mean length of broad cervices (category 3) wasn’t significantly different from with those with normal size with associated pathology (p = 0.37). This is because only two among eight patients in category three had normal AP diameter of cervices despite having an associated pathology. Thus, it can be assumed that the presence of a pathology is often associated with enlargement of uterine cervices.

Analysis of the subgroup with pair of ovaries containing developing follicles without any pathology (n=27) revealed moderate correlation (figure 1f) between the ovarian volumes (R = 0.53, p = 0.004) with the mean (±SD) ovarian volume as, 6.73±3.6 cc on left and 8.54 ± 5.2 cc on right. In this subgroup, the mean(±SD) of difference in the ovarian volume was 1.81±4.4 cc, where the left ovaries were significantly smaller than those on right (figure 4a, t-test p-value = 0.04). The right ovary was smaller than the left in eight (29.6%) patients with mean (± SD) of difference -3.3±3.1 (figure 4b). The right ovary was larger than the left in 19 (70.4%) patients with mean (± SD) of difference being 3.95 ± 2.9 cc (figure 4b).

**Figure 4.**
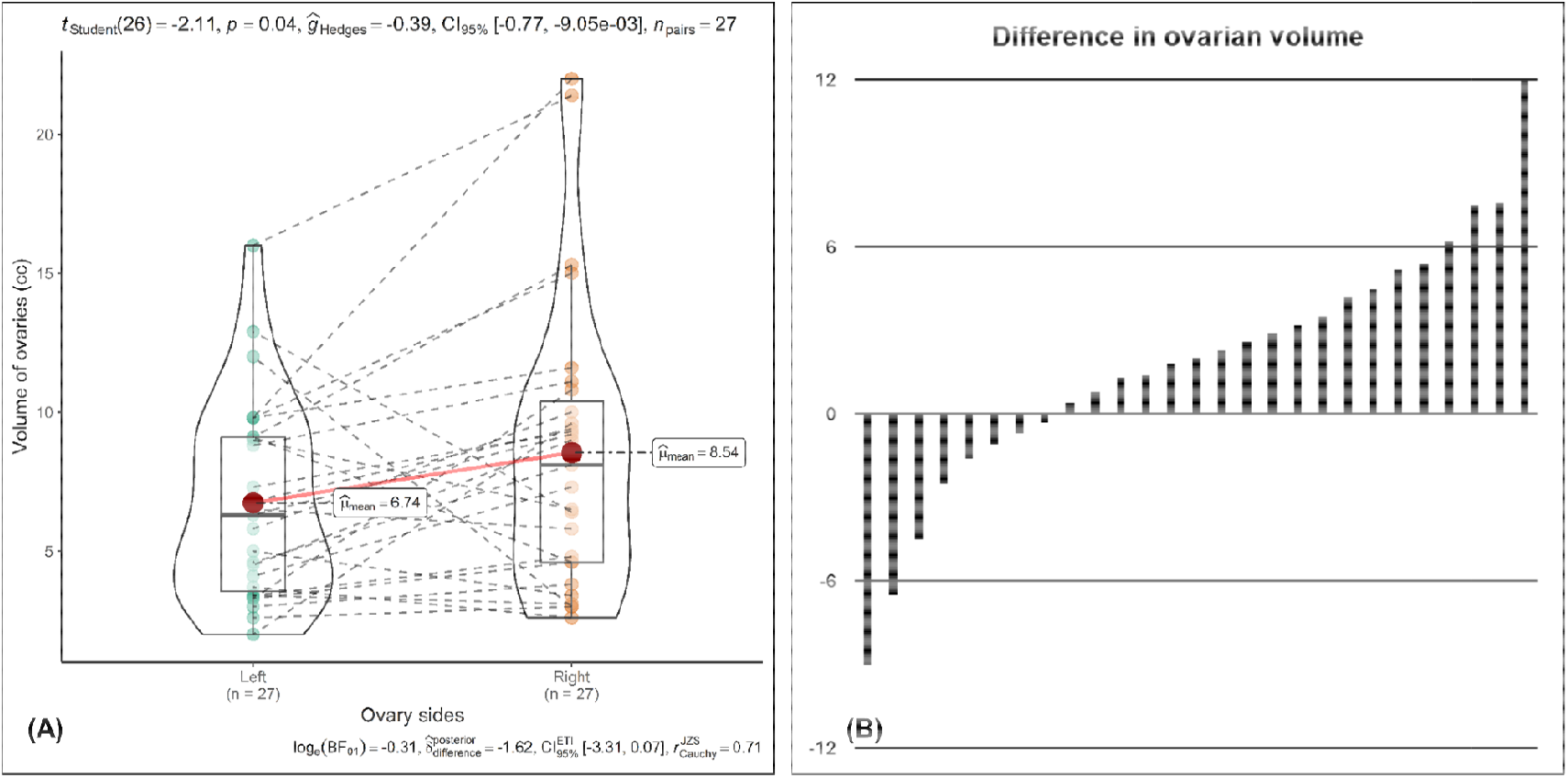
Box-violin plot (A) showing distribution of ovarian volume in pair with no associated pathology. The left ovarian volume was overall smaller than that of the left. Waterfall plot (B) with the difference of volume (right ovary – left ovary) plotted in Y-axis shows, the right ovary was smaller than the left in eight (29.6%) patients and larger than the left in 19 (70.4%) patients.

## DISCUSSION

The patients with bulky uterus in this series were more likely to have myoma or adenomyosis in comparison to those having normal sized uterus. Sonographic findings of bulky uterus along with altered myometrial echotexture is considered to have a diagnostic performance that is nearly equivalent to that of a pelvic MRI (9, 10). Among all the parameters observed in this study, only the fundal AP diameter of uterus showed significant correlation with patients’ age while the other uterine and cervical length and diameters, despite no significant correlation to age (data not shown), showed, correlation with length of uterus. Additionally, the previously reported correlations of uterine AP diameter (11, 12) are poor in comparison to the current study

While the patients’ age is reportedly in a positive correlation with ovarian volumes in pre-pubertal girls (11, 13, 14) there’s an observed negative correlation in the perimenopausal women (15-17) with, reportedly smaller ovarian volume among users of hormonal contraceptives (18, 19). This complex interaction is a reason for non-discovery of a direct correlation between age and ovarian volume in this series (data not shown) comprising of small number of patients.

The reported reference values for ultrasound organometry from the western world are often adopted as the local standard for practice of ultrasound. However, there remain concerns about those reference values not being suitable for a proportion of the local population.

Surprisingly, we observed normal ovaries with volume of up to 22 cc which is comparable to the reported upper limit of 20 cc from a large cohort (20). The ovarian asymmetry reported in this current series is a documentation of fact that often goes underestimated in the daily practice of ultrasound. More research in this area is expected to widen the current understanding with potential implication on management of infertility. Furthermore, development of a centile chart for the pelvic organs from local population data is expected to have clinical and forensic implications.

## CONCLUSION

This study is a documentation of facts from routine practice of TVS that were compared to the reported facts to understand the difference of our data as well as to explain the characteristic of our data. The distribution of uterus and cervix size across finding-based categories and the volume asymmetry of normal ovaries presented herein will contribute to a better understanding of the morphological variations and pathological features of the uterus, cervix and ovaries, potentially paving some future research with clinical and forensic implications.

## Data Availability

All data produced in the present study are available upon reasonable request to the corresponding author

